# Informal payments in health facilities in Peru in 2018: analysis of a cross-sectional survey

**DOI:** 10.1101/2023.03.28.23287883

**Authors:** Laura A. Espinoza, Patricia Mallma, Hannah H. Leslie, Patricia J. García

## Abstract

**Background:** The Latin American region demonstrates the lowest levels of trust in health systems globally. Institutional corruption is a major factor in eroding trust. Corruption in health services, including extracting bribes and informal payments from patients, directly harms health outcomes and weakens services intended as public goods. In this study, we aim to characterize the frequency and distribution of informal payments within public health services in Peru.

**Methods:** We conducted a secondary analysis of a nationally representative cross-sectional survey, the 2018 National Household Survey of Living Conditions and Poverty and identified all individuals reporting health insurance from the Ministry of Health (SIS-MINSA) or Social Security (ESSALUD). To assessed self-reported informal payments in the population, we defined it in 2 ways: 1) being asked to pay a bribe at a health establishment in the past year (direct method), and 2) non-zero cost of care for services that should be free (indirect method) in the last month. We used descriptive statistics to quantify informal payments and bivariate analysis to identify sociodemographic characteristics of those most frequently reporting such payments.

**Findings:** 132,355 people were surveyed, including 69,839 (52.8%) with coverage from SIS-MINSA and 30,461 (23.03%) from ESSALUD. 39% of respondents reported informal payments in public health services. Less than 1% of participants directly reported them, either at SIS-MINSA services (0.22%); or at ESSALUD (0.42%). Indirect reporting was more prevalent, including up to 10% of surgery patients and 17% of those hospitalized in SIS-MINSA facilities. Wealthier patients (19%) were more likely to report such payments. Interpretation: While direct reporting of bribery was uncommon, we found moderate prevalence of informal payments in public health services in Peru using an indirect assessment method. Indirect reporting may exceed direct reporting due to difficulty in distinguishing appropriate and inappropriate payments, fear of reporting health care workers’ behavior, or social tolerance of informal payments. Informal payments were more common among those with greater social and financial capital, indicating they may obtain enhanced services. Further research on patients’ perception and reporting of informal payments is a key step towards accurate measurement and evidence-based intervention.

## Introduction

Corruption affects health systems around the world (1–5). According to García (6), the health sector is an attractive sector for corruption due to its complexity and the asymmetry of power. Corruption in service delivery can be classified in six common types: absenteeism, informal payments, embezzlement during service provision, favoritism, and manipulation of data. In addition, corruption is one of the most important obstacles worldwide because many of its forms, such as the diversion of funds, bribes as economic barriers to care, health personnel who do not attend at the established hours and do not provide the care that the population should receive, make it much more difficult to implement universal health coverage (7). Corruption in health systems limits access to health services and weakens all the dimensions that determine the different domains of quality of health care: limiting access, undermining quality, and worsening inequity, each of which impedes health systems from delivering on the promise of better health for all (8) Corruption in the health sector is not only a legal or economic phenomenon, but a social phenomenon: a set of practices that are embedded and normalized in the routine experience of health service users (9,10). Poor oversight systems and weak accountability mechanisms are associated with greater corruption in health systems. (9)

Informal payments in health are a type of corruption (11). While multiple definitions of such payments have been proposed, a systematic review (12) recommends, “a direct contribution, which is made in addition to any contribution determined by the terms of entitlement, in case or in-kind, by patients or others acting on their behalf, to health care providers for services that the patients are entitled to” (3) as the most clear and neutral definition. Operationalizing services to which patients are entitled in a specific health system is essential to capturing informal payments.

Informal payments in healthcare can occur in different situations within the user’s care process. For example, when patients may pay to shorten waiting time or bypass queues at services, to obtain medicines or medical supplies in short supply, or to accelerate access to surgical services. In some cases, those payments take the form of gifts, or non-monetary goods, which may come from people who are not necessarily sick, but who need other types of services such as falsification of medical documents (13) (14) (15).

Important repercussions on the socioeconomic aspect have been given by the prevalence of informal payments, as well as on the accessibility of health services, especially for the population living in vulnerable situations. As a matter of fact, a prior study in 33 low-income countries in Africa found informal payments were important to advantage those with more resources and hence exacerbate disparities in access to health care between poor and rich as well as disparities between regions (16).

Measurement and evaluation of the prevalence associated factors and main services in the health system which present informal payments is an important step in the search for improving the quality of health care for users and in the search and work for equity and effective compliance. The way that informal payments have been defined and measured is not standardized; measurement approaches include both qualitative and quantitative methods, particularly household survey records and corruption surveys. For instance, in Peru, a postgraduate study by the Universidad Peruana Cayetano Heredia conducted in 2018 estimated the prevalence of informal payments in users of the public health insurance from the Ministry of Health (SIS-MINSA) from 2008 to 2010 using information collected in the National Household Survey of Living Conditions and Poverty (ENAHO). This study found a prevalence of expenditures outside the tariff framework and during the health care process in all 25 regions of the country. It was also shown that the frequency of SIS-MINSA users who made these payments at some point during their process of care increased from 27% in 2008 to almost 35% in 2010. (17)

Health coverage in Peru has changed since 2010: SIS-MINSA health system was expanded to fully subsidizes the provision of services not only to school children, mothers, and newborns, but to the population living in poverty and extreme poverty who come mostly from rural and marginal urban areas. In addition, the instrument used for recollecting information, *ENAHO* survey structure, has changed adding modules particularly focused on the evaluation of the population’s experience with government/public entities to assess governance directly asking for bribes or illegal situations.

The objective of this study is to describe prevalence of informal payments in public health services in Peru following the expansion of public health coverage, identifying types of health services and individuals most prone to informal payments.

## Methods

### Study setting

Peru’s health system has two sectors, public and private. The public sector is divided into the subsidized regime (*Seguro Integral de Salud*, SIS-MINSA) and the direct contributive regime or social security (ESSALUD).

Since 2010, to guarantee the full right to health, the Universal Health Coverage Law states that the entire population in the territory must have health insurance that allows and cover access to the list of insurable conditions, which includes preventive, promotional, recuperative and rehabilitation services.

### Data source

The present study carried out a secondary analysis of the information of the Opinion Module and the Health Module of the ENAHO 2018 cross-sectional survey. This survey considers as a target population all private dwellings and household members, divided into three groups according to the level of relationship: head of household (over 18 years old), spouse (over 12 years old) and other members, residing in rural and urban areas at the national level. For the purposes of the study, we will consider the same target population of the ENAHO, but who are affiliated to the Comprehensive Health Insurance (SIS-MINSA) or Social Security (ESSALUD). The annual size of the ENAHO 2018 sample defined by the National Institute of Statistics and Informatics of Peru (*INEI* in Spanish). (https://m.inei.gob.pe/) is 39 820 individual households, corresponding to 24 308 households in urban areas and 15 512 households in rural areas. The sample of clusters at the national level is 5 752, corresponding to 3 813 clusters in urban areas and 1 939 clusters in rural areas.

### Measures

There is no single gold standard measure for informal payments. (12) We used two complementary methods of assessment included in the ENAHO questionnaire to measure informal payments in Peru. The first was the direct method, which consisted of reporting being asked to pay a bribe at a public health establishment in the past year. This question was asked only of participants who were over 18 years of age and received care in any SIS-MINSA or ESSALUD facility in the last 12 months. Respondents were asked if they had been asked for a bribe or gift and separately if they had given one. We considered answering yes to either question as an indicator of informal payments, and we classified within this outcome whether payments had been solicited and/or provided.

The second one was the indirect method; we calculated reporting any type of informal payment or bribe among all affiliates of SIS-MINSA and ESSALUD who used health services within the past 12 months. Since recall periods were shorter for services like medication use, this indicator may underestimate informal payments on an annual basis. Services included on the questionnaire were: medicines in the last 4 weeks; glasses and contraception in the last 3 months; inpatient attention and surgical intervention in the last year. Based on national policy in 2018, these services should be provided without charge to SIS-MINSA and ESSALUD affiliates.

The independent variables were sociodemographic characteristics of the population: sex, age, degree of education, level of poverty (INEI 2018 classification: population whose monthly expenditure is less than the cost of a basic food and non-food consumption basket [approx. 95 USD], considered poor; and the population whose per capita expenditure does not cover the cost of the basic food consumption basket [approx. 50 USD], considered extremely poor), geographical stratum and marital status.

### Statistical analysis

We used descriptive statistics to describe the sample of health service users within the government’s services and to report prevalence of informal payments using Chi Square test. Then, we conducted a bivariate analysis using Fisher’s exact test and linear regression to compare frequency of informal payments for each health service type across sociodemographic characteristics. We conducted this analysis with the goal of describing those groups most affected by informal payments and selected unadjusted comparisons as the best approach to meeting this goal.

The sample design involves up to three sampling stages where units are selected with probabilities proportional to size except for the last stage. In the last stage, several dwellings are selected for each cluster considering a selection interval. Moreover, the expansion factors are adjusted considering the population projections by age and sex groups for each survey month and inference levels proposed in the sample design. Thus, all analyses in this study were conducted using sample strata and survey weights.

We used the statistical program STATA version 17.0 for the analysis.

### Ethics Statement

This research project was approved by the Institutional Ethics Committee of the Universidad Peruana Cayetano Heredia under code SIDISI 201297 in January 2020. Verbal consent was obtained as part of the original study from ENAHO 2018. Ethical review of this secondary analysis of deidentified data confirmed no further consent was required.

Due to this is a cross-sectional study, the work was reported following the requirements of the STROBE guidelines and the checklist is in Appendix 1.

## Results

Of the 39,820 households sampled, ENAHO surveyed 132,355 people. Within them, 69,839 (52.80%) were affiliated to SIS-MINSA and 30,461 (23.03%) to ESSALUD (Table 1). SIS-MINSA patients have a higher proportion of poor persons with respect to ESSALUD (32.56% vs 5.71% in poverty, p<0.001), and a higher percentage come from rural areas (36.80% MINSA vs. 4.65% ESSALUD, p<0.001). In addition, ESSALUD patients have a higher proportion of affiliates with undergraduate education (36.23% ESSALUD vs. 9.53% MINSA p<0.001).

**Table 1.**
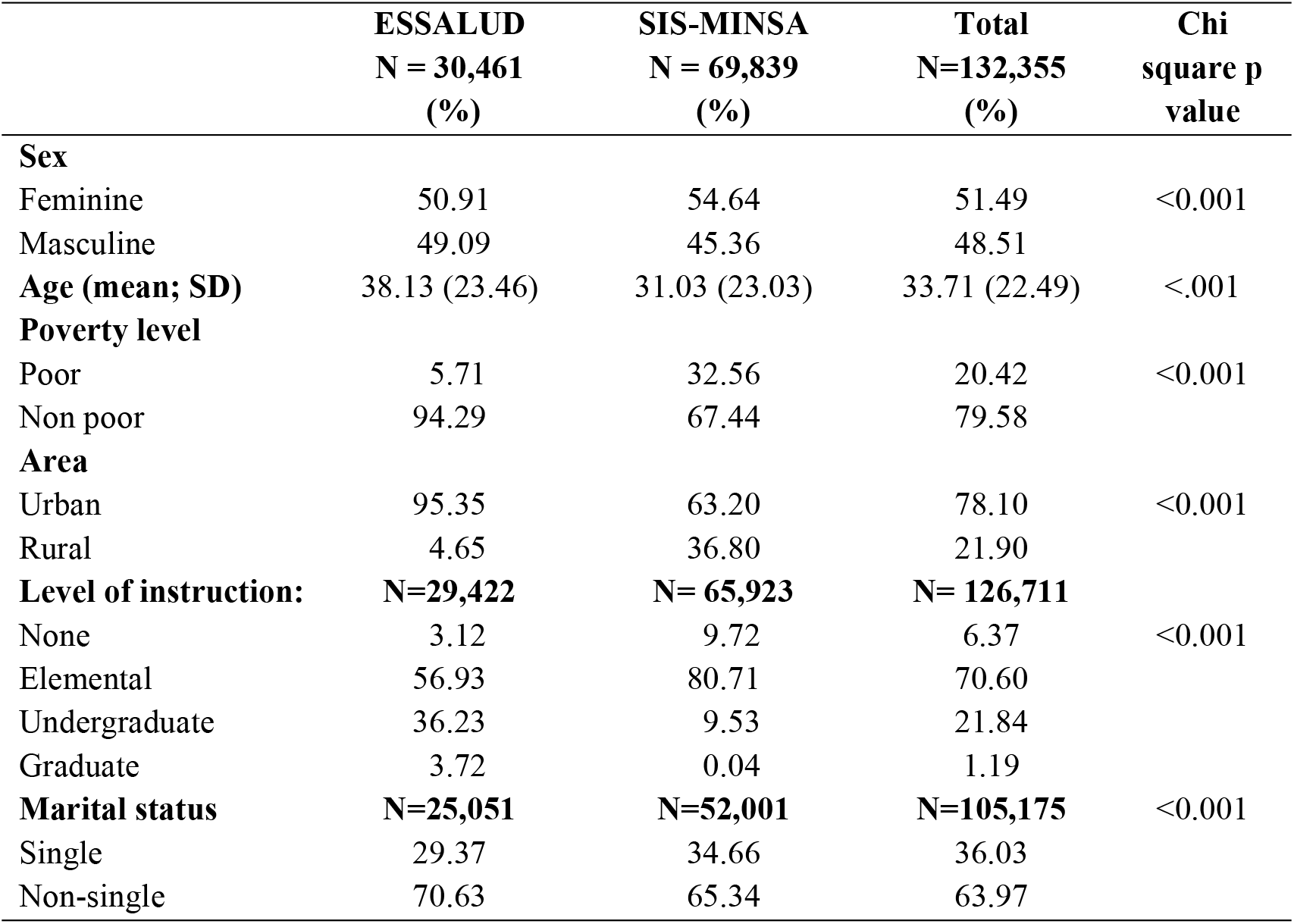
Demographics characteristics of ESSALUD and SIS-MINSA user’s.

Eligible insurance participants for analysis this prevalence in the last year approached by the direct method, were 3,964 in ESSALUD and 11,458 in SIS-MINSA. The prevalence of informal payments was 0.42% ESSALUD and 0.22% SIS-MINSA (Fig. 1). In the case of ESSALUD, 0.23% of the affiliates were asked, given; 0.33 were asked, not given; 0.19% were not asked, given and 99.25% were not asked, not given informal payments. On the other hand, within SIS-MINSA, 0.14% were asked, given; 0.50 were asked, not given; 0.08 were not asked, given and 99.28% were not asked, not given.

**Fig 1.** Prevalence of informal payments in ESSALUD and SIS-MINSA - direct method approach

By the indirect method, the highest prevalence of informal payments made for users in the last year in SIS-MINSA services were 17.37% during in-patient services and 10.86% during surgical intervention (Fig.2). In the case of ESSALUD services, payments were made in 4.11% of in-patient services and 1.11% of surgical interventions.

**Fig 2.** Informal payments prevalence in ESSALUD and SIS-MINSA health services - indirect method approach

Given the low prevalence of directly reported informal payments and some services with indirectly reported informal payments, we focused on characterizing prevalence higher than 4%. According to the bivariate analysis of the sociodemographic characteristic among SIS-MINSA users, informal payments were significantly more commonly reported by non-poor respondents (19% vs. 13.3%) and urban (19.2% vs. 15.8%) within those using inpatient services (Table 2). No significant differences were identified for ESSALUD respondents.

**Table 2.** Sociodemographic characteristics of population in high-reported informal payments health services.

|  | In-patient |  |  |  |  | Surgical intervention |  |  |  |
| --- | --- | --- | --- | --- | --- | --- | --- | --- | --- |
|  | ESSALUD<br>N = 1,793<br>(%) |  | Fisher exact<br>test p<br>value | MINSA<br>N = 3,042<br>(%) |  | Fisher exact<br>test p<br>value | MINSA<br>N = 1,539<br>(%) |  | Fisher exact<br>test p<br>value |
|  | Si | No |  | Si | No |  | Si | No |  |
| **Sex** |  |  | 0.709 |  |  | 0.136 |  |  | 0.290 |
| Feminine | 3.83 | 96.17 |  | 17.16 | 82.84 |  | 10.66 | 89.34 |  |
| Masculine | 4.17 | 95.83 |  | 19.42 | 80.58 |  | 8.93 | 91.07 |  |
| **Level of instruction** |  |  | 0.944 |  |  | 0.086 |  |  | 0.056 |
| None | 3.77 | 96.23 |  | 14.34 | 85.66 |  | 10.74 | 89.26 |  |
| Basic | 3.76 | 96.24 |  | 18.44 | 81.56 |  | 8.54 | 91.46 |  |
| Undergrad | 4.29 | 95.71 |  | 18.60 | 81.40 |  | 14.29 | 85.71 |  |
| Graduate | 3.90 | 96.10 |  | 100 | 0.00 |  | 0.00 | 100 |  |
| **Poverty level** |  |  | 0.720 |  |  | **0.001\*** |  |  | 0.196 |
| Poor | 1.96 | 98.04 |  | 13.33 | 86.67 |  | 7.17 | 92.83 |  |
| Non-poor | 4.02 | 95.98 |  | 19.07 | 80.93 |  | 9.87 | 90.13 |  |
| **Stratum** |  |  | 0.845 |  |  | **0.016\*** |  |  | 0.050 |
| Urban area | 3.94 | 96.06 |  | 19.27 | 80.73 |  | 10.63 | 89.37 |  |
| Rural area | 4.17 | 95.83 |  | 15.88 | 84.12 |  | 7.63 | 92.37 |  |
| **Marital status** |  |  | 0.588 |  |  | 0.330 |  |  | 0.132 |
| Single | 4.60 | 95.40 |  | 19.72 | 80.28 |  | 12.21 | 87.79 |  |
| Non-single | 3.84 | 96.04 |  | 17.79 | 82.21 |  | 9.05 | 90.95 |  |
|  |  |  | **Linear regression p value** |  |  | **Linear regression p value** |  |  | **Linear regression p value** |
| **Age (mean; SD)** |  |  | 0.780 |  |  | 0.868 |  |  | 0.205 |
|  | 42.63<br>(24.67) | 43.78<br>(24.09) |  | 35.43<br>(21.22) | 35.20<br>(22.73) |  | 35.99<br>(16.87) | 38.70<br>(20.54) |  |

## Discussion

The present study analyzed the prevalence of informal payments with direct question and an indirect question about informal payments in different health services for participants who reported having public insurance from either ESSALUD or SIS-MINSA. We found that, when asked directly, the prevalence of informal payments was very low (0.42% ESSALUD and 0.22% SIS-MINSA). However, when patients were approached indirectly, the reporting percentages were much higher, especially in the in-patient service (MINSA: 17.37%, ESSALUD: 4.11%) and surgical intervention (MINSA: 10.86%; ESSALUD: 1.11%). In addition, there were no significant differences in sociodemographic characteristics for ESSALUD patients who reported informal payments and just a few for SIS-MINSA. In the case of the last one, individuals with more social capital were more likely to report such payments.

According to the literature, the informal payments system could lead to “redistribution” among users, with welfare/administrative staff playing a “Robin Hood” role, subsidizing the poor and charging the rich (16). One of the reasons for this to happen could be that health personnel represent a “collection agency for medical charities” that solicits the richest above marginal cost and uses the proceeds to provide care for the poorest. However, none of these studies has been able to verify that this situation really benefits the poor insured with respect to access, quality, and satisfaction of care. Another important reason could be that poor users would not be able to make informal payments due to their economic limitations, so the same health personnel do not address them with those kinds of questions.

The areas with the greatest presence of informal payments are the in-patient area from both SIS-MINSA and ESSALUD; and surgical intervention area from SIS-MINSA, this could be justified because nature of the services in the public health systems. There are more saturated because there are no methods of help for timely diagnosis, medical care instruments, equipment of different complexity, and the deficiency in the process of acquiring medicines and medical supplies for these services (18,19). Here, informal payments could be in cash or in another way to pay different members of the medical staff involved in the patient care process within the in-patient or surgery area: the attending physician, the surgeon, the anesthesiologist, the nurses, among others. While there is no evidence patients in Peru must buy their own supplies, this has been demonstrated in other countries in transition, receiving medical treatment during in-patient service will require patients to contribute the necessary care materials, including medicines, medical supplies, and even non-medical supplies. Therefore, when a person is hospitalized, relatives may supplement the resources of the available health centers by providing some kind of nursing service regarding the patient, in addition to being in charge of the medications and injections that correspond to him (20).

Within the same national representative survey, individuals who were directly asked about informal payments reported a much lower frequency compared to those who only indicated that they had made payments while insured. A challenge in operationalizing informal payments is clarity on the benefits to which the patient is entitled free of charge (12). Differentiating between informal payments and out-of-pockets payments may be difficult due to a lack of patients’ awareness of their legal rights and coverage entitlements. In that sense, it may be valid to assume that all transactions where patients report being not given any information about official co-payments rates or fees are informal. (21) (5) (22) Future efforts to measure informal payments may consider assessment of respondents’ awareness of entitlements and expected payments.

A greater rate of informal payments in the same resource according to the indirect approach may be due to the social desirability bias in national surveys that seek to record citizen behavior, in a context such as Peru, where corruption is widespread and social norms may conflict when asking about corrupt behavior or complicity in corrupt behavior (23). In this sense, it’s suggested to generate data by selecting other strategies that reduce the discomfort of respondents when having to answer a sensitive question. It could be achieved using alternatives approaches of recollection of informal situations like the indirect method for informal payments, which take into consideration participants who attended public services and requested services which are free and fully covered by law but are willing to pay to obtain free access to full healthcare coverage and medications. Items that do not directly mention “informal”, “illegal” or “corruption” reduce the potential risk-perception to the respondents.

Considering just the indirect reporting of informal payments, our results suggest a decline in prevalence compared to the analysis of the 2010 national study (17). Although both analyze informal payments with almost the same set of questions from the cross-sectional national survey ENAHO, the structure in 2010 and 2018 version has changed. Likewise, comparing results over time is not completely feasible when expansion of social security and the broader availability of affordable services have been implemented in the public health system. These conditions could explain the differences in lower reporting.

Since this is a secondary analysis, we used the questions that had already been asked, and adapted the variables to define informal payments. We cannot fully delimit who the informal payments were made (administrative staff of the institution or health service providers, for example). Similarly, due to the type of survey used, it is not known when the informal payments was conducted or if there was coercion. (18) We do not have information on the amounts in the transaction of these informal payments, which could also allow us to estimate the losses they represent and the out-of-pocket expense for the insured. We have evaluated one side of the problem, however, it would be ideal to know the attitude of health personnel in these cases, as well as the motivations they could have as a result of receiving or requesting informal payments.

Finally, the main strength of our study is the update of the analysis of the prevalence of informal payments collected annually in SIS-MINSA and ESSALUD establishments since 2010. The present study complemented two analytical strategies for measuring informal payments. In addition, having analyzed the results of ENAHO, it allows us to make informal payments calculations that can be approximated at the national level. Furthermore, it allows us to identify the ways that the government develops ways of measuring sensitive topic as corruption. Patients may not be aware of the benefits they are entitled to increasing communication around covered services and assessing individuals’ awareness of such coverage is a strategy to consider. Although this information is available online, there are additional strategies could be considered for better outreach. As is mentioned in the literature, the surveys that addressed informal payments should collect data by appropriate data collection strategies. More accurate measurements and further research is necessary in order to report the real estimate of informal payments. This study’s results are of interest and useful to share with government health entities towards the generation of evidence-based actions for the benefit of the population.

## Data Availability

The present study carried out a secondary analysis of the information of the Opinion Module and the Health Module of the ENAHO 2018 cross-sectional survey. This survey considers as a target population all private dwellings and household members, divided into three groups according to the level of relationship: head of household (over 18 years old), spouse (over 12 years old) and other members, residing in rural and urban areas at the national level. For the purposes of the study, we will consider the same target population of the ENAHO, but who are affiliated to the Comprehensive Health Insurance (SIS-MINSA) or Social Security (ESSALUD). The annual size of the ENAHO 2018 sample defined by the National Institute of Statistics and Informatics of Peru (INEI in Spanish). (https://m.inei.gob.pe/) is 39 820 individual households, corresponding to 24 308 households in urban areas and 15 512 households in rural areas. The sample of clusters at the national level is 5 752, corresponding to 3 813 clusters in urban areas and 1 939 clusters in rural areas.

## Appendix 1

STROBE Statement—Checklist of items that should be included in reports of ***cross-sectional studies***.

## References

1. Gaal P, McKee M. Informal payment for health care and the theory of “INXIT.” Int J Health Plann Manage. 2004;19(2):163–78.

2. Jowett M, Danielyan E. Is there a role for user charges? Thoughts on health system reform in Armenia. Bulletin of the World Health Organization. 2010 Jun 1;88(6):472–3.

3. Gaal P, Evetovits T, McKee M. Informal payment for health care: evidence from Hungary. Health Policy. 2006 Jun;77(1):86–102.

4. Mæstad O, Mwisongo A. Informal payments and the quality of health care: Mechanisms revealed by Tanzanian health workers. Health Policy. 2011 Feb;99(2):107–15.

5. Belli P, Gotsadze G, Shahriari H. Out-of-pocket and informal payments in health sector: evidence from Georgia. Health Policy. 2004 Oct;70(1):109–23.

6. García PJ. Corruption in global health: the open secret. Lancet. 2019 Dec 7;394(10214):2119–24.

7. Transparency International. Annual Report 2006 - Publications [Internet]. Transparency.org. 2006 a[cited 2022 Nov 24]. Available from: https://www.transparency.org/en/publications/transparencyinternational-annual-report-2006

8. Kruk ME, Gage AD, Arsenault C, Jordan K, Leslie HH, Roder-DeWan S, et al. High-quality health systems in the Sustainable Development Goals era: time for a revolution. The Lancet Global Health. 2018 Nov 1;6(11):e1196–252.

9. Mujica J. Huber, Ludwig. La corrupción como fenómeno social. Romper la mano. Una interpretación cultural de la corrupción. Lima: Proética, Instituto de Estudios Peruanos, 2008, 179 pp. Anthropologica. 2008 Mar 29;26(26):247–51.

10. Mackey TK, Vian T, Kohler J. The sustainable development goals as a framework to combat health-sector corruption. Bull World Health Organ. 2018 Sep 1;96(9):634–43.

11. Lewis M. Informal payments and the financing of health care in developing and transition countries. Health Aff (Millwood). 2007;26(4):984–97.

12. Cherecheş RM, Ungureanu MI, Sandu P, Rus IA. Defining informal payments in healthcare: A systematic review. Health Policy. 2013 May 1;110(2):105–14.

13. World Health Organization. Informe sobre la salud en el mundo: la financiación de los sistemas de salud: el camino hacia la cobertura universal [Internet]. Organización Mundial de la Salud; 2010 [cited 2022 Nov 24]. Available from: https://apps.who.int/iris/handle/10665/44373

14. Burki T. Corruption is an “ignored pandemic.” The Lancet Infectious Diseases. 2019 May 1;19(5):471.

15. World Health Organization. Human rights and health [Internet]. 2017 [cited 2022 Nov 24]. Available from: https://www.who.int/news-room/fact-sheets/detail/human-rights-and-health

16. Kankeu HT, Ventelou B. Socioeconomic inequalities in informal payments for health care: An assessment of the “Robin Hood” hypothesis in 33 African countries. Soc Sci Med. 2016 Feb;151:173–86.

17. Rosas Febres ME. Inequidad dentro de la equidad pagos informales durante la atención de los afiliados al Seguro Integral de Salud 2008 a 2010 [Internet]. Universidad Peruana Cayetano Heredia; 2018 [cited 2022 Nov 24]. Available from: https://repositorio.upch.edu.pe/handle/20.500.12866/5953

18. Soto A. Barreras para una atención eficaz en los hospitales de referencia del Ministerio de Salud del Perú: atendiendo pacientes en el siglo XXI con recursos del siglo XX. Revista Peruana de Medicina Experimental y Salud Publica. 2019 Jun;36(2):304–11.

19. Gutiérrez C, Romaní Romaní F, Wong P, Del Carmen Sara J. Brecha entre cobertura poblacional y prestacional en salud: un reto para la reforma de salud en el Perú. Anales de la Facultad de Medicina. 2018 Jan;79(1):65–70.

20. World Health Organization. Regional Office for Europe, Policies EO on HS and, Kutzin J, Cashin C, Jakab M. Implementing health financing reform: lessons from countries in transition [Internet]. World Health Organization. Regional Office for Europe; 2010 [cited 2022 Dec 15]. 411 p. Available from: https://apps.who.int/iris/handle/10665/326420

21. Mirabedini SA, Fazl Hashemi SME, Sarabi Asiabar A, Rezapour A, Azami-Aghdash S, Hosseini Amnab Hassan. Out-of-pocket and informal payments in Iran’s health care system: A systematic review and meta-analysis. Med J Islam Repub Iran. 2017 Oct 5;31:70.

22. Hernández-Vásquez A, Rojas-Roque C, Vargas-Fernández R, Rosselli D. Measuring Out-of-pocket Payment, Catastrophic Health Expenditure and the Related Socioeconomic Inequality in Peru: A Comparison Between 2008 and 2017. J Prev Med Public Health. 2020 Jun 10;53(4):266– 74.

23. Baraybar Hidalgo V, Guibert Y, Muñoz P. Bribing and Social Desirability in Peru: A Mixed Methods Approach. Colombia Internacional. 2022 Apr 1;(110):21–49.

